# SARS-CoV-2 Antigen Rapid Detection Tests: test performance during the COVID-19 pandemic and the impact of COVID-19 vaccination

**DOI:** 10.1101/2024.04.11.24304791

**Authors:** Isabell Wagenhäuser, Kerstin Knies, Tamara Pscheidl, Michael Eisenmann, Sven Flemming, Nils Petri, Miriam McDonogh, Agmal Scherzad, Daniel Zeller, Anja Gesierich, Anna Katharina Seitz, Regina Taurines, Ralf-Ingo Ernestus, Johannes Forster, Dirk Weismann, Benedikt Weißbrich, Johannes Liese, Christoph Härtel, Oliver Kurzai, Lars Dölken, Alexander Gabel, Manuel Krone

## Abstract

**Introduction:** During the COVID-19 pandemic, SARS-CoV-2 antigen rapid detection tests (RDTs) emerged as point-of-care diagnostics in addition to the RT-qPCR as the gold standard for SARS-CoV-2 diagnostics. Facing the course of the COVID-19 pandemic to an endemic characterised by several SARS-CoV-2 virus variants of concern (VOC) and an increasing public COVID-19 vaccination rate the aim of the study was to investigate the long-term test performance of SARS-CoV-2 RDT in large-scale, clinical screening use during and its influencing factors, above all SARS-CoV-2 VOC and COVID-19 vaccination.

**Methods:** In a prospective performance assessment conducted at a single centre tertiary care hospital, RDTs from three manufacturers (NADAL®, Panbio™, MEDsan®) were compared to RT-qPCR among individuals aged ≥ 6 month. The evaluation involved the determination of standardised viral load from oropharyngeal swabs as well as the evaluation of their influencing factors, especially the COVID-19 vaccination, for detecting SARS-CoV-2 in a clinical point-of-care environment spanning from 12 November 2020 to 30 June 2023 among patients, staff, and visitors of the hospital.

**Results:** Among the 78,798 RDT/RT-qPCR tandems analysed, 2,016 (2.6%) tandems tested positive for SARS-CoV-2, with an overall sensitivity of 34.5% (95% CI 32.4-36.6%). A logistic regression revealed that typical COVID-19 symptoms significantly declined over the course of the study and throughout the COVID-19 pandemic, and that among the vaccinated, significantly fewer presented with an infection exhibiting typical symptoms. The employed lasso regression model indicated that only higher viral load and typical COVID-19 symptoms significantly increase the likelihood of a positive RDT result in the case of a SARS-CoV-2 infection directly.

**Conclusion:** Our findings indicate that only viral load and COVID-19 symptoms directly influence RDT performance while the obtained effects of COVID-19 vaccination and Omicron VOC both reducing RDT performance were mediated by these two factors. RDTs remain an adequate diagnostic tool for detecting SARS-CoV-2 in individuals showing respiratory symptoms. RDTs show promise beyond SARS-CoV-2, proving adaptable for detecting other pathogens like Influenza and RSV, highlighting their ongoing importance in infection control and prevention efforts.

## 1 Introduction

Since 2020, the COVID-19 pandemic has posed far-reaching global challenges,[1] new therapies,[2] prevention strategies, and diagnostic concepts have been developed in a very short space of time and repeatedly adapted to and influenced by the dynamics of the pandemic.[3]

The acute phase of the pandemic has now passed, and COVID-19 is in transition as a seasonal pathogen of acute respiratory diseases and is now a new player alongside Influenza and RSV.[4–8]

In the acute phase of the COVID-19 pandemic in particular, an important pandemic management strategy, alongside the development of therapies and vaccines and the establishment of public measures such as contact restrictions, was the timely, rapid and reliable diagnosis of SARS-CoV-2 as the central key to breaking chains of infection.[9–12]

As a well-established, very precise method, reverse transcription polymerase chain reaction (RT-qPCR) has been the gold standard for diagnostics since the beginning of the pandemic.[9] For a more rapid, cost-effective and point-of-care diagnostics, SARS-CoV-2 rapid tests (RDT) were made available as lateral flow immunoassays just a few months after the beginning of the pandemic, with cost-effective point-of-care application without infrastructural requirements and rapid results.[13]

To date, a large body of evidence has demonstrated in detail that the sensitivity and specificity of RDTs can be far below the manufacturer’s specifications and do not correspond to the gold standard of RT-qPCR although the majority of the evidence to date does not cover the entire COVID-19 pandemic, analyses only small, selective test collectives or does not evaluate RDTs in screening use.[14] The following correlations have already been proven in the evidence to date as decisive factors influencing test performance: the presence of typical COVID-19 symptoms and high viral load correlate positively with high sensitivity values of RDTs.[13, 15–21]

However, since the establishment of RDTs in COVID-19 diagnostics, many circumstances in the test environment have changed which requires the re-evaluation of RDT performance under this current conditions.[22] With the course of the pandemic, the infestation of society and the availability of COVID-19 vaccines, there is now a basic immunised test collective.[23, 24] As the various SARS-CoV-2 virus variants of concern (VOC) progressed, the initial wild-type SARS-CoV-2 was chronologically displaced first by the Alpha and Delta VOC and ultimately by the Omicron VOC with its various sublines, which paved the way from pandemic to endemic with lower morbidity and a population that was immunised in parallel by previous infections and the available vaccinations.[25, 26]

Evidence to date is heterogeneous that the Omicron VOC might reduce the test performance of RDTs.[16, 19, 21, 27–30] Data from an interim analysis of the study suggested that any deterioration in RDT test performance is not attributable to VOC itself but rather to the change in symptomatology mediated by the VOC throughout the course of the COVID-19 pandemic.[18] Further, regarding the potential influence of COVID-19 vaccination on RDT performance only very few studies have so far considered the aspect of COVID-19 vaccination revealing no influence of COVID-19 vaccination on RDT performance.[19, 29, 31, 32] A preprint from 2022, not yet published in the peer-review process, discussed the hypothesis based on their results that the observed decrease in RDT sensitivity in clinical use, despite higher viral loads, is attributable to increased immunity among the study population due to COVID-19 vaccinations and previous SARS-CoV-2 infections.[32] In contrast, the two previously published studies that consider the potential influence of COVID-19 vaccination status on the large-scale clinical RDT test performance factor do not observe any impact of vaccination status on RDT performance. However, they only cover the pandemic period up to early 2022. [19, 29]

This is the first study analysing the large-scale test performance and its influencing factors of RDTs in clinical screening use, including the role of COVID-19 vaccination and SARS-CoV-2 VOC, in the longitudinal course of the COVID-19 pandemic until its endemic transition in 2023.

## 2 Methods

### 2.1 Test enrolment

As part of the strategies implemented to prevent and mitigate the spread of SARS-CoV-2 within the hospital setting in the beginning of the COVID-19 pandemic, a tandem RDT/RT-qPCR testing was employed at a German 1,438-bed tertiary care hospital from 12 November 2020 to hinder the transmission of nosocomial SARS-CoV-2 chains starting.

From 12 November 2020 to 24 November 2022, mandatory entry screening, i.e., RDT diagnostics upon admission, was conducted for patients and inpatient companions in all critically assessed areas of the hospital, such as emergency departments and delivery rooms, based on the prevailing evidence and pandemic situation. During periods of high COVID-19 incidence, from 1 February 2021, to 30 June 2021, and from 4 November 2021, to 24 November 2022, this mandatory entry screening was extended to all areas of the hospital, resulting in a universal RDT entry screening.[23, 25]

In addition, since 12 November 2020, employees with COVID-19 typical symptoms or contact with a SARS-CoV-2 positive person were examined using RDT/RT-qPCR test tandem at the central testing center of the hospital. The documented RDT/RT-qPCR tandems were also included in the study.

From 25 November 2022, mandatory entry screening at the hospital, involving both RDT and RT-qPCR, was discontinued due to the general easing of the pandemic situation, and the sole use of RT-qPCR continued. The use of additional RDTs or RDTs without parallel RT-qPCR for screening was subsequently implemented and documented in risk-adapted, decentral concepts customised for their individual characteristics.

On 19 May 2023, with the complete transition from pandemic to endemic, this cross-clinic, individualised mandatory RDT deployment strategy and further diagnostic continuation were voluntary in individual clinics until 30 June 2023, which remarks the end of the study period (*Supplementary Figure 1*).[23, 25, 33]

### 2.2 Data collection

The following **inclusion criteria** were defined for considering a paired RDT / RT-qPCR result for the analysis:

- documented RDT with parallel RT-qPCR
- valid test result of the RDT (presence of a control line, no interference lines)
- age ≥ six month

This age limit was deliberately chosen against the background of the EMA’s vaccine authorisations for individuals aged six months and older in the course of the study in order to be able to analyse the influence of COVID-19 vaccination on RDT performance.[34–37]

Documented RDTs were excluded from data analysis in the following situations:

- multiple testing (more than one RDT per day and person): only the first chronologically performed RDT per day and person were considered. Patients meeting the inclusion criteria on multiple days during the study period underwent testing and inclusion once per visit.
- recent SARS-CoV-2 infections and subsequent deisolation were excluded from the analysis due to the potential persistence of RT-qPCR positivity unrelated to the risk of viral transmission.[38]

The sampling for RDT/RT-qPCR test tandems was consistently carried out by trained healthcare workers, ensuring correct execution. The swabs were taken as paired, consecutively collected oropharyngeal samples.

The overall dataset was merged from the following sources (*Figure 1*).

- hospital information system (HIS; SAP ERP 6.0 (SAP, Walldorf, Germany)): RDT documentation, RT-qPCR results, demographic data, clinical information, and anamnestic information on COVID-19 vaccination
- hospital’s COVID-database with a systematic overview about all positive SARS-CoV-2 detections at the hospital
- epidemiological data on VOC prevalence in Germany[25]
- standardised viral load calculation of the RT-qPCR positive samples as described below
- EMA COVID-19 vaccination authorisation data

**Figure 1:**
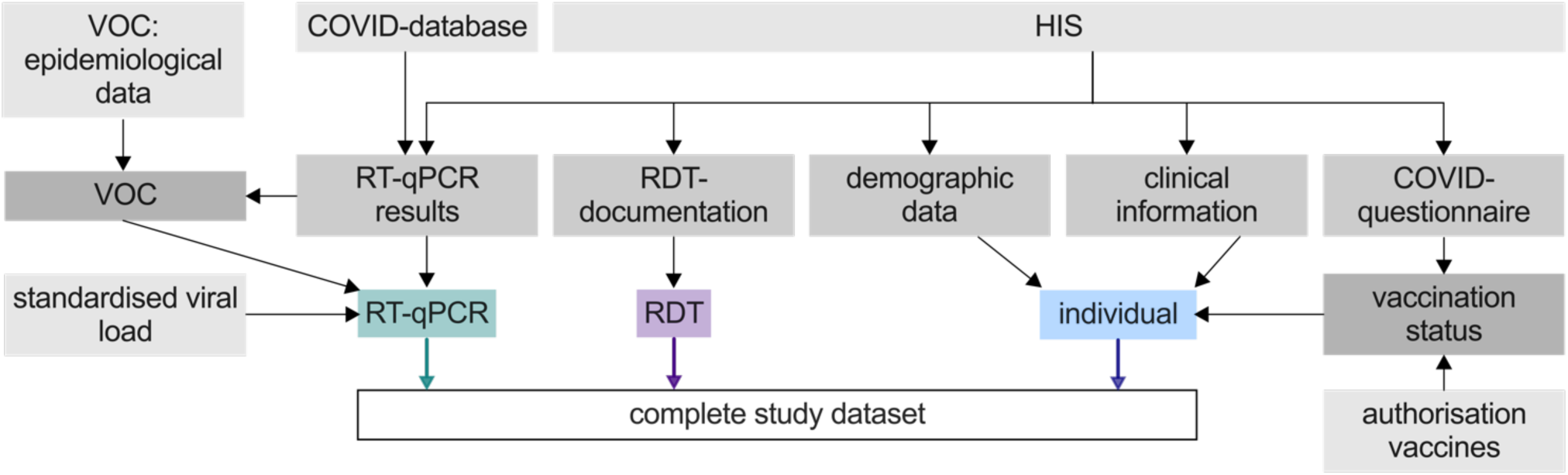
Schematic overview of data acquisition. RDT: Antigen rapid detection test RT-qPCR: Quantitative reverse transcription-polymerase chain reaction VOC: SARS-CoV-2 virus variant of concern HIS: hospital information system

Subjects were classified based COVID-19 case definition provided by the CDC[39] and the ECDC[40] into the following cohorts:

- **typical COVID-19 symptoms**: individuals suffering fever, dry cough, shortness of breath, new anosmia, or ageusia
- **atypical COVID-19 symptoms** potentially be linked to COVID-19: individuals with a decline in general condition, falls, diarrhoea, or seizures
- **asymptomatic** individuals

The vaccination status of the patients at the time of each RDT was determined by evaluating the admission questionnaire and incorporating the official approval data of the COVID-19 vaccines by the European Medicines Agency (EMA). The COVID-19 questionnaire was conducted as part of the standardised entry interview at the study centre from 31 May 2021, for the entire study period recording whether the patient was immunised against COVID-19 with at least two doses of an EU-approved COVID-19 vaccine or with at least one dose in addition to a confirmed PCR-confirmed SARS-CoV-2 infection. Until the most recent documentation of the vaccination status “unvaccinated” of a subject, all this and previously conducted RDTs were classified as RDTs with as “unvaccinated”. From the first documentation of a vaccination onwards, all RDTs conducted for an individual were classified as conducted with a fully “vaccinated” COVID-19 immune status. In addition, all RDTs performed before the age-stratified initial approval of a COVID-19 vaccine by the EMA were classified as having an “unvaccinated” COVID-19 immune status. In the age group with a minimum age of 16 years, all conducted RDTs before 21 December 2020, were classified as “unvaccinated”,[34] in the age group between 12 and 15 years all RDTs before 28 May 2021,[35] in the age group between five and 11 years all RDTs before 26 November 2021,[36] and in the age group between four years and six month all RDTs before 19 October 2022.[37]

### 2.3 Antigen rapid detection tests (RDT)

To maintain an uninterrupted logistical provision, three specific RDT were chosen from a pool of 23 products identified by the German Federal Institute for Drugs and Medical Devices in October 2020.[15, 41] All the RDTs used are listed on the EU Common List of COVID-19 antigen tests by the European commission (directorate-general for health and food safety).[42]

I. NADAL® COVID-19 Ag Test (Nal von Minden GmbH, Regensburg, Germany)
II. Panbio™ COVID-19 Ag Rapid Test (Abbott Laboratories, Abbott Park IL, USA)
III. MEDsan® SARS-Cov-2 Antigen Rapid Test (MEDsan GmbH, Hamburg, Germany)

All three RDTs used for the study are designed as lateral flow immunoassays with the SARS-CoV-2 nucleoprotein antigen as the target structure, according to manufacturer information. NADAL® and MEDsan® RDTs are approved for use with oropharyngeal swabs. The Panbio™ RDT is approved for nasopharyngeal swabs, but in this study, it was also used with oropharyngeal swabs in comparison to RT-qPCR.

The distribution of RDTs to the individual hospital’s departments was random, depending on availability, independent of the current RDT deployment concept. All RDTs performed as part of the study were carried out directly at the point of care, decentralised immediately after the swab, following manufacturer instructions by trained medical personnel, and results were documented. Since RT-qPCR diagnostics were only available after the RDT processing time due to logistics and RT-qPCR processing time, the interpretation of the RDT was always done without knowledge of the RT-qPCR result.

### 2.3 RT-qPCR and viral load determination

RT-qPCR diagnostics were processed in the hospitals’ virological diagnostic laboratories utilising several RT-qPCR methods adhering to the guidelines provided by the respective manufacturers.

To prioritise RDT-positive samples for the fastest possible confirmation by RT-qPCR, the RDT results were made available to the virus diagnostics staff.

The subsequent RT-qPCR analytical instruments were employed for the determination of viral load:

I. MagNaPure 96 / 7500 Real-Time PCR System / FTD SARS-CoV-2-PCR (target N/ORF1ab-gene, Roche Diagnostics, Rotkreuz, Switzerland / Thermo Fisher Scientific, Waltham MA, USA / Siemens Healthineers, Munich, Germany)
II. NeuMoDx™ (target N/Nsp2-gene, Qiagen, Hilden, Germany)
III. Alinity m (target RdRp/N-gene, Abbott Laboratories, Abbott Park IL, USA)
IV. QIAstat-Dx® (target RdRp/E-gene, Qiagen)
V. Xpert® Xpress SARS-CoV-2/Flu/RSV (target E/N2/RdRp-gene, Cepheid, Sunnyvale CA, USA)
VI. cobas® SARS-CoV-2 (target ORF1ab/E-gene, Roche Diagnostics)
VII. cobas® Liat (target ORF1a/b/N-Gen, Roche Diagnostics)
VIII. BIOFIRE® FILMARRAY® (target S-/M-gene, bioMérieux, Marcy-l’Étoile, France)

Since the two methods cobas® Liat and BIOFIRE® FILMARRAY® only enable qualitative SARS-CoV-2 RT-qPCR without quantification of a Cycle Threshold value (C_t_-value), in the event of positive SARS-CoV-2 detection by one of these two methods, the RT-qPCR was repeated for quantification with Xpert® Xpress SARS-CoV-2/Flu/RSV in case of a positive cobas® Liat result and Xpert® Xpress SARS-CoV-2/Flu/RSV or NeuMoDx™ in case of a positive BIOFIRE® FILMARRAY® result.

Viral loads were computed from C_t_-values employing the previously described formula with reference standards, as follows:[15]

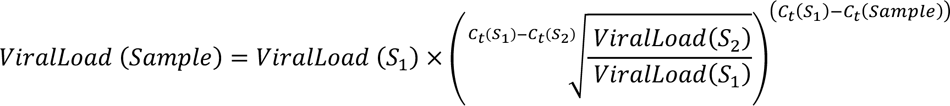

For instances involving multiple targets with distinct C_t_-values on an RT-qPCR system (cobas®, NeuMoDx™, Xpert® Xpress SARS-CoV-2/Flu/RSV), the viral load was determined by computing the geometric mean of the estimates derived from the two individual genes (*Supplementary Figure 2*).

### 2.4 SARS-CoV-2 virus variant of concern

Between February 3, 2021, and January 19, 2022, for allocation of VOC of all RT-qPCR-positive samples with sufficient viral load a PCR with spike protein variant-specific differentiation was performed. Outside of the phase of molecular VOC determination, variant assignment was done epidemiologically wherever possible. The precise procedure of molecular and epidemiological VOC assignment is described in the *Supplementary Methods* and *Supplementary Table 1*.[23, 25, 43]

### 2.5 Ethical approval

The Ethics committee of the University of Würzburg considered the study protocol and waived the need to formally apply for ethical clearance due to the study design (File Nr 20231219 02).

### 2.6 Statistics

The data in the overall RDT dataset were recorded using Excel 2019 (Microsoft, Redmond WA, USA). The hospital’s COVID-19 database is based on an Access 2019 (Microsoft, Redmond WA, USA) platform. Statistical analyses were conducted using GraphPad Prism 10.2.1 (GraphPad Software, San Diego CA, USA), and R (Version 4.1.3).

Confidence intervals were calculated using the Wilson-Brown method (RDT test performance) or the Baptista-Pike method (Odds Ratio).[44]

Statistical significance levels were calculated using the Fisher-Exact test (comparison of sensitivity by manufacturers, VOC, vaccination status, and symptoms) and the Mann-Whitney U-test (comparison of viral loads).

Regression analyses were employed to investigate the influence of the following factors on viral load (linear regression analysis) and typical COVID-19 symptoms (logistic regression analysis): age, gender, COVID-19 vaccination, and testing time point during the COVID-19 pandemic.

A logistic Lasso regression was performed to identify factors associated with the RDT result confirming a SARS-CoV-2 infection. The regression model included factors age, gender, viral load typical COVID-19 symptoms, COVID-19 vaccination, and infection by the Omicron VOC. Using a tenfold cross-validation procedure, the model parameters of the Lasso regression model were estimated (*Supplementary Figure 3*).To correct against multiple testing, the resulting p-values were adjusted using the Benjamini-Yekutieli procedure.[45] .

For both regression analyses, only those RDT/RT-qPCR tandems with available vaccination status as well as a clearly epidemiologically or molecularly assigned VOC were considered.

Adjusted p-values < 0.05 were considered statistically significant.

## 3 Results

### 3.1 Test enrolment

Between 12 November 2020 and 30 June 2023, a total of 113,117 RDTs were performed and documented at the study centre from individuals aged ≥ 6 month. After exclusion of RDTs without parallel RT-qPCR, multiple RDTs on one study day and in case of a recent de-isolation as well as RDTs with invalid results, 78,798 RDT/RT-qPCR test tandems from 53,918 individuals could be included (*Figure 2*). The 48 invalid RDTs are distributed among the three RDT manufacturers as follows: 5 (10.4%) NADAL®, 15 (31.2%) PANBIO™ and 28 (58.3%) MEDsan®.

**Figure 2:**
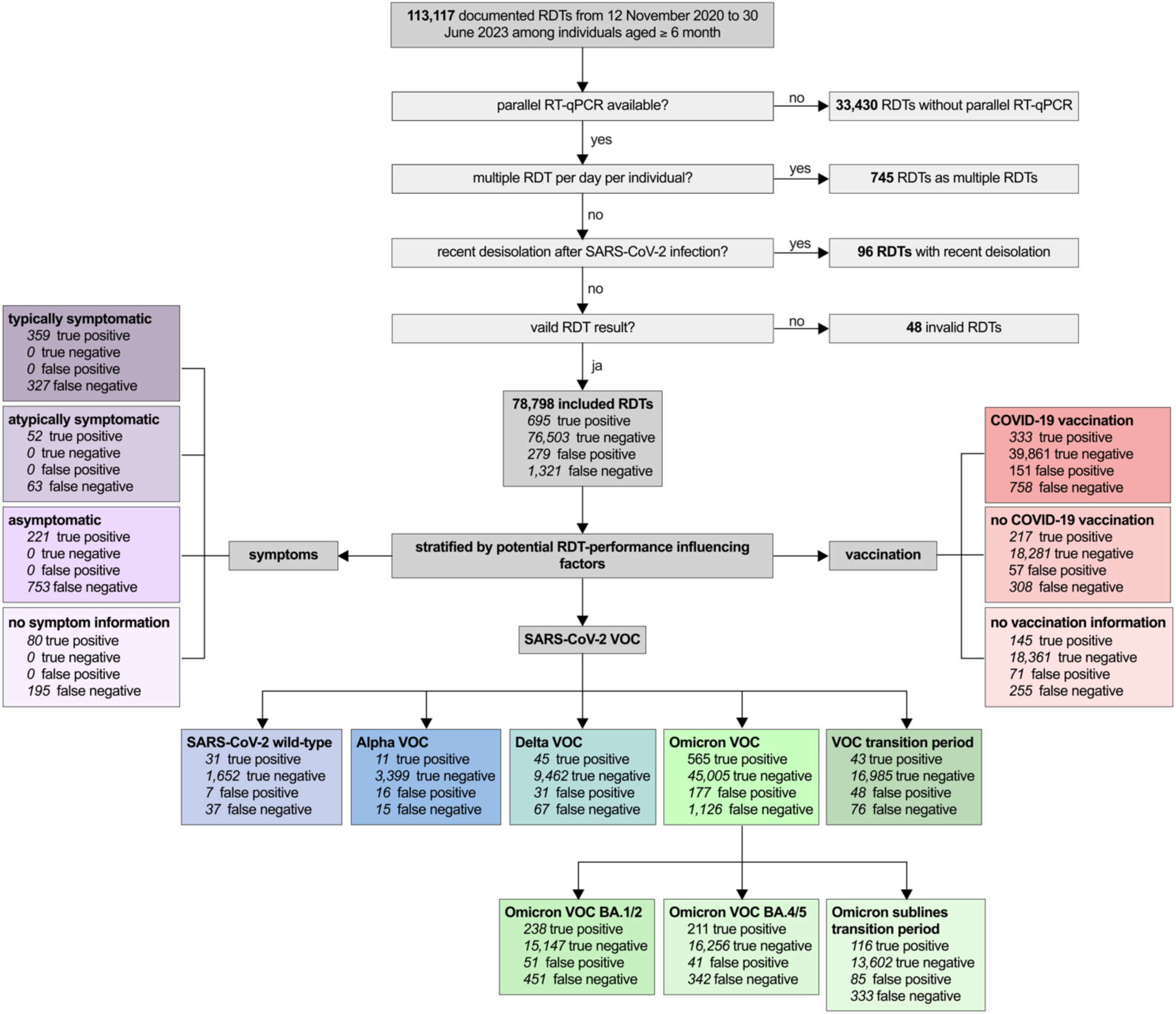
Enrolment of SARS-CoV-2 antigen rapid detection tests (RDTs) VOC: virus variant of concern RDT: Antigen Rapid Detection Test RT-qPCR: Quantitative reverse transcription-polymerase chain reaction

### 3.2 Study population

The median **age** of the individuals included at paired RDT/RT-qPCR performance analysis was 54 years (range 6 month to 102 years, IQR: 31-70 years) covering 49.5% (39,037/78,798) female and 50.5% (39,759/78,798) male individuals (*Figure 4A*). Two RDT/RT-qPCR test tandems were performed on individuals allocating themselves to diverse **gender**.

The RDT/RT-qPCR test tandems included were performed in 87.3% (68,819/78,798) on patients, in 11.7% (9,228/78,798) on accompanying individuals, and in 1.0% (751/78,798) on staff.

Overall, a **SARS-CoV-2 prevalence** of 2.6% (2,016/78,798) was detected in the entire cohort (*Figure 3A*) with a median viral load of 5.6 (IQR: 4.1-7.2) log_10_ SARS-CoV-2 RNA copies/ml (*Supplementary Figure 5*). In the linear regression analysis, the factors age (p=0.40), gender (p=0.94), timing of RDT/RT-qPCR tandem tests during the study (p=0.16), and COVID-19 vaccination (p=0.44) did not show any significant association with viral load.

**Figure 3:**
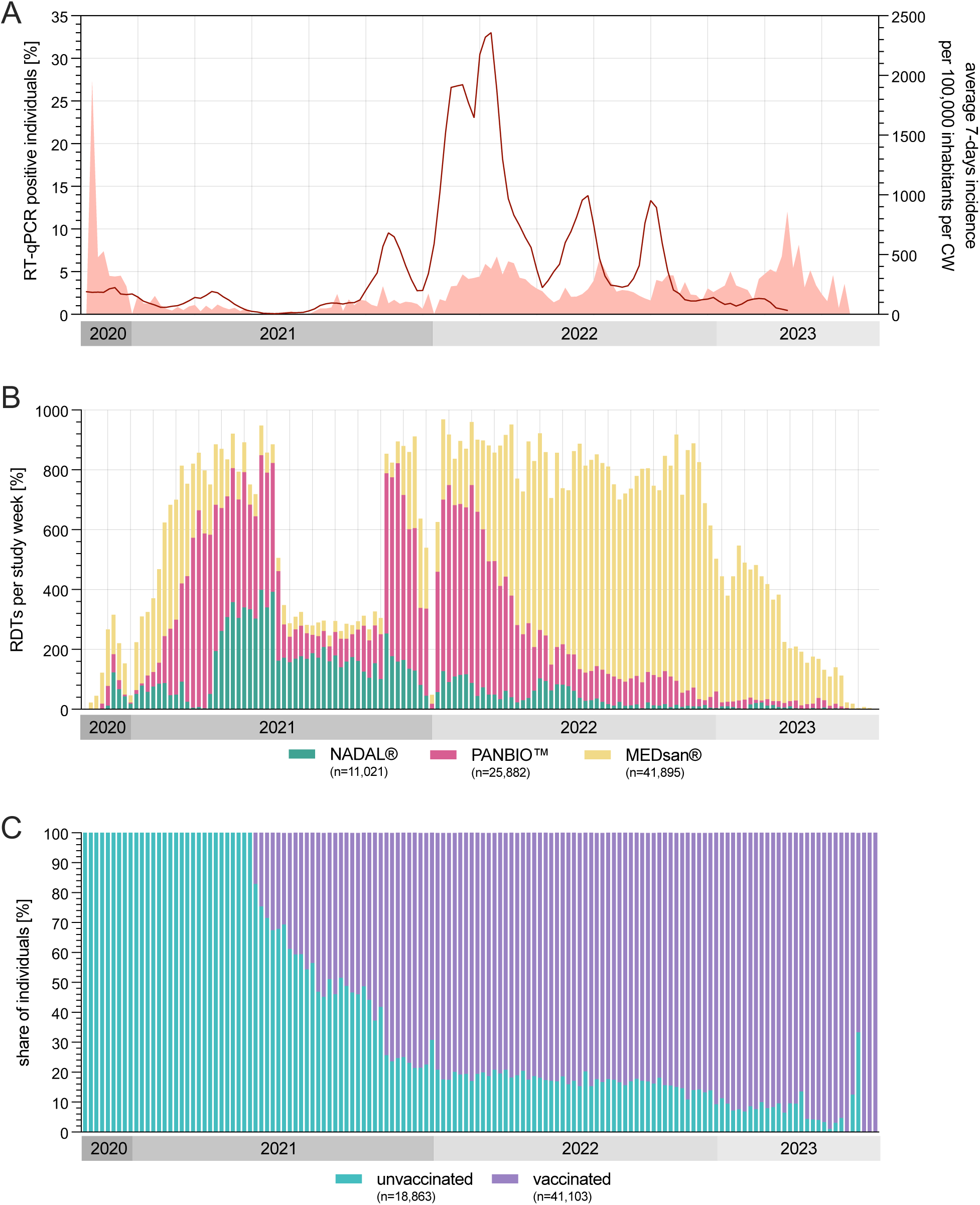
SARS-CoV-2 prevalence, number of tests, and COVID-19 vaccination status over the course of the study. 3A) Fraction of RT-qPCR positive RDT/RT-qPCR test tandems per study week (bright red filled curve) and SARS-CoV-2 incidence per 100,000 inhabitants per CW in the study region (red line) 3B) Number of RDTs per CW stratified by RDT manufacturer 3C) Fraction of COVID-19 vaccination status of RDT/RT-qPCR test tandems per CW CW: Calendar week RDT: Antigen Rapid Detection Test RT-qPCR: Quantitative reverse transcription-polymerase chain reaction Data source: Bayerisches Landesamt für Gesundheit und Lebensmittelsicherheit[23, 25]

Regarding the **manufacturer** 11,021 (14.0%) of the test tandems were performed with NADAL®, 25,882 (32.8%) with PANBIO™ and 41,895 (53.2%) with MEDsan® (*Figure 3B*).

Among the test tandems with positive RT-qPCR results, 3.4% (68/2,016) could be allocated to wild-type SARS-CoV-2, 1.3% (26/2,016) to the Alpha **VOC**, 5.6% (112/2,016) to the Delta VOC, 34.2% (689/2,016) to the Omicron BA.1/2 VOC and 27.4% (553/2,016) to the Omicron BA.4/5 VOC. The remaining 29.1% (586/2,016) could not be allocated to any VOC whereas 76.6% (449/586) were in the transition phase between the two Omicron VOC groups. Two (0.1%; 2/2,016) further RDT/RT-qPCR samples were molecularly allocated to the Iota VOC; in one (0.1%; 1/2,016) remaining sample no VOC could be finally allocated despite molecular testing.

Information on **COVID-19 vaccination status** was available among 76.1% (59,966/78,798) of included RDT-/RT-qPCR samples, where of 31.5% (18,863/59,966) were conducted on unvaccinated and 68.5% (41,103/59,966) on vaccinated individuals (*Figure 3C*). Among the remaining 23.9% (18,832/78,798) of individuals no information on COVID-19 vaccination status could be evaluated (*Supplementary Figure 4*). A progressive proportion of the included RDT/RT-qPCR tandems on vaccinated individuals were chronologically performed from the Delta VOC period onwards (*Figure 4B*).

Among the RT-qPCR positive tandems, 34.0% (686/2,016) presented with COVID-19 typical and 5.7% 115/2,016) with COVID-19 atypical **symptoms**. 48.3% (974/2,016) were asymptomatic (*Figure 4, Supplementary Figure 5*). In 315 (45.9%) of the typically symptomatic test tandems included, information on the number of days since symptom onset was available. The viral load decreased significantly in the disease course (p<0.0001; *Figure 4C*). The proportion of individuals tested SARS-CoV-2 positive with COVID-19 symptoms increased by viral load, decreased in the chronological sequence of the VOC periods and was lower among vaccinated individuals (*Figure 4*). A logistic regression analysis demonstrated a significant decrease in typical COVID-19 symptoms over the study period (p=0.021). Moreover, among those who were COVID-19 vaccinated, there was a significant reduction in the number of individuals experiencing a SARS-CoV-2 infection with typical symptoms (p<0.0001). The factors of age (p=0.16) and gender (p=0.83) showed no significant influence on typical symptomatology.

**Figure 4:**
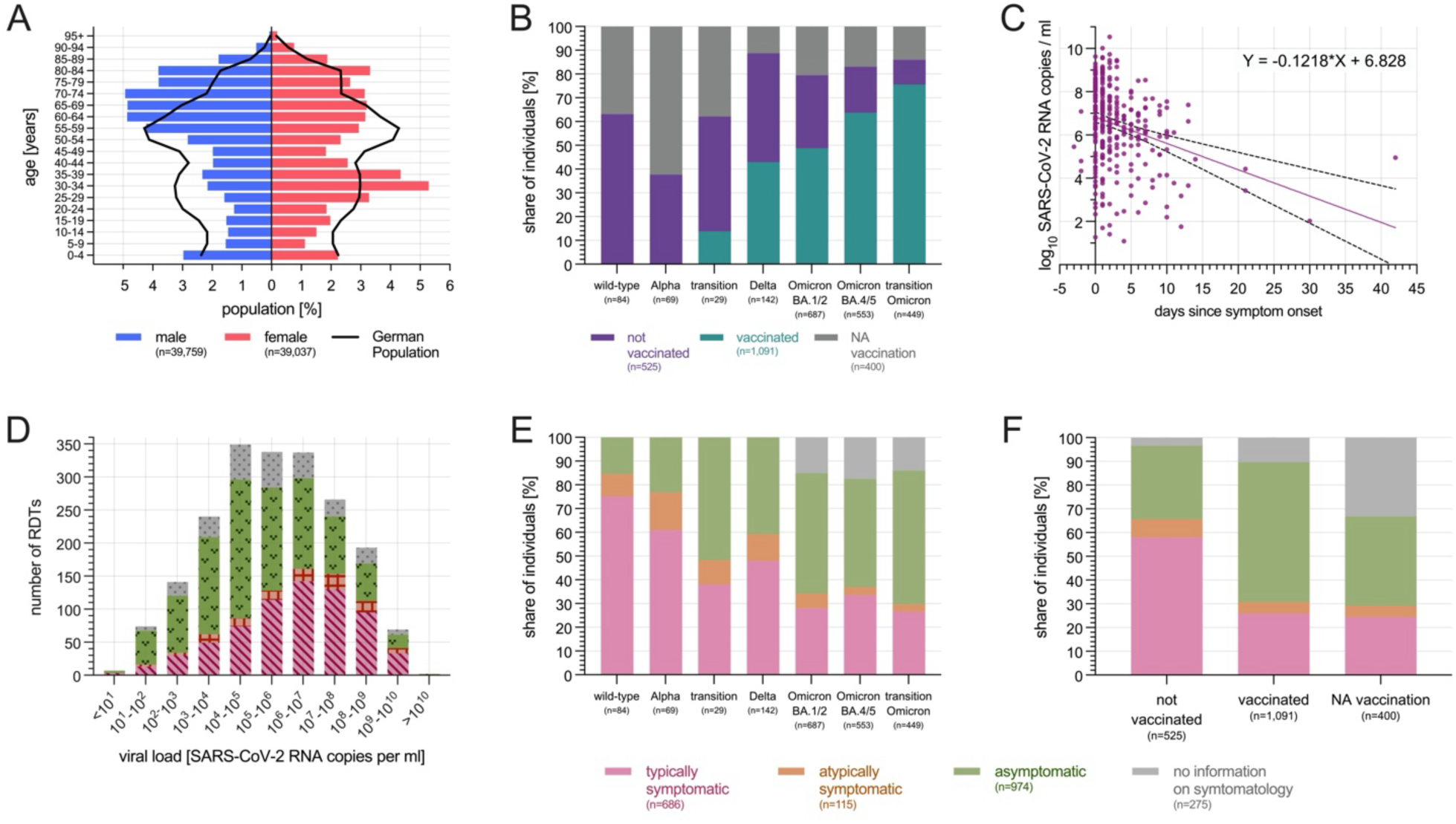
Characterisation of the study population. 4A) Age characterisation in categories of five years stratified by gender in reference to the German population 4B) COVID-19 vaccination status stratified by SARS-CoV-2 VOC among RT-qPCR positive samples 4C) Viral load depending on days since symptom onset 4D) Distribution of viral load stratified by symptoms in absolute numbers among RT-qPCR positive samples 4E) COVID-19 symptom status stratified by SARS-CoV-2 VOC among RT-qPCR positive samples 4F) COVID-19 symptom status stratified by vaccination status among RT-qPCR positive samples RDT: Antigen Rapid Detection Test NA: no information available Data source: Bavarian State Office for Statistics[46]

### 3.3 RDT performance compared to RT-qPCR: univariate analyses

Overall, a RDT **sensitivity** of 34.5% (95% CI 32.4-36.6%) and a RDT specificity of 99.6 (95% CI 99.6-99.7%) was obtained (*Figure 5A*). Regarding sensitivity no differences between the different manufacturers (all p>0.40) and the different SARS-CoV-2 VOC (all p>0.14) could be obtained in the univariate comparison. Univariately, the sensitivity among asymptomatic individuals was significantly lower compared to typically as well as atypically symptomatic individuals (both p<0.0001). Further the sensitivity among vaccinated individuals was significantly lower compared to the unvaccinated (p<0.0001; *Figure 5A*).

**Figure 5:**
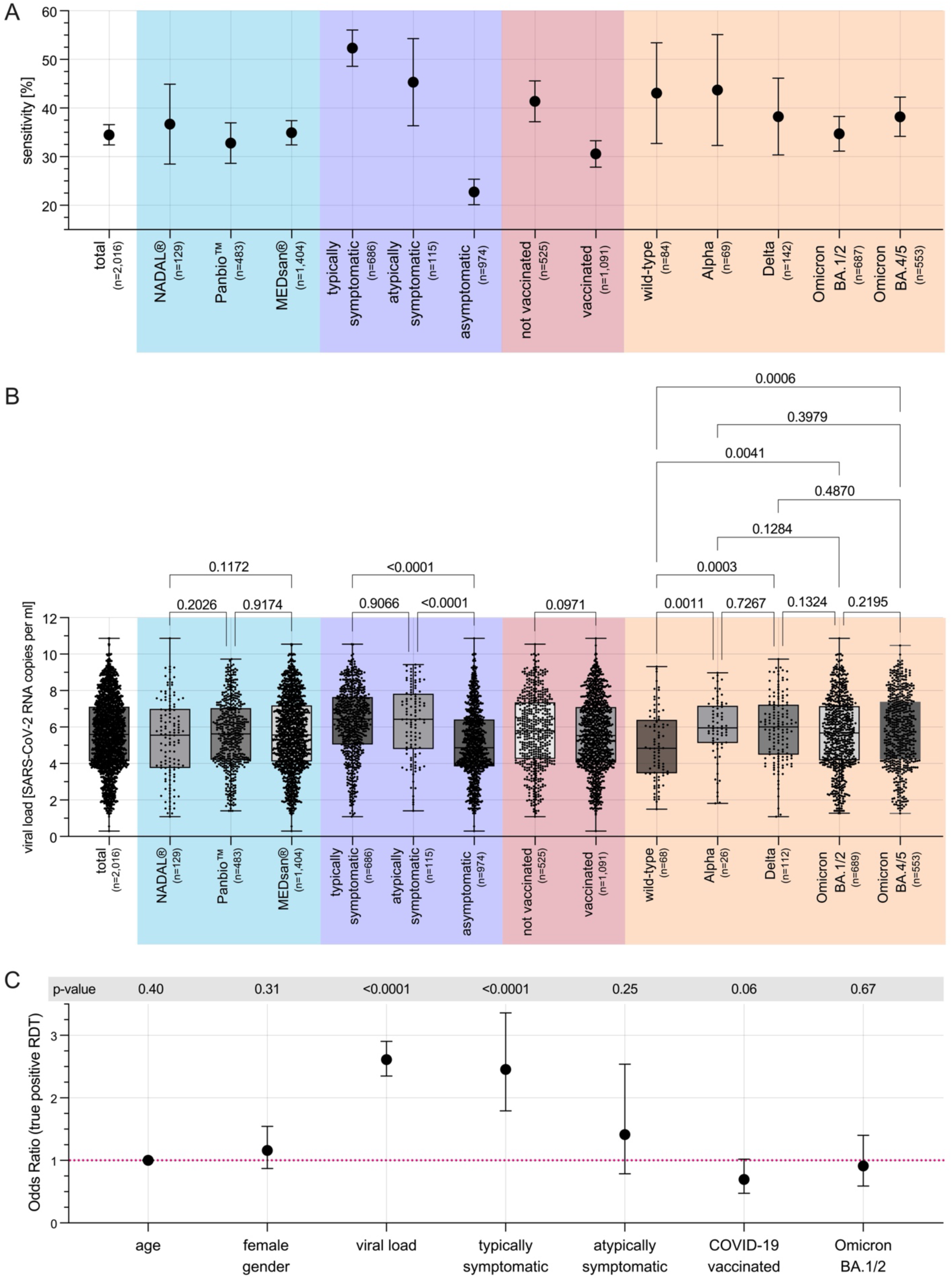
SARS-CoV-2 prevalence, number of tests, and COVID-19 vaccination status over the course of the study. 5A) RDT sensitivity overall and by RDT performance influencing factors 5B) Viral load overall and by RDT performance influencing factors 5C) Odds Ratio of the several RDT performance influencing factors (lasso regression model) In the case of whiskers in the figures, these represent the respective 95% confidence intervals. VOC: virus variant of concern RDT: Antigen Rapid Detection Test

Regarding the **viral load**, no significant differences could be obtained comparing univariately comparing the different manufactures (all p>0.12), as well as comparing vaccinated to unvaccinated individuals (p=0.10). Regarding the SARS-CoV-2 VOC the mean viral load of wild-type samples was significantly lower comparing to Alpha VOC samples (p=0.0011), to Delta VOC samples (p=0.0003), to Omicron BA.1/2 VOC samples (p=0.0041) and to Omicron BA.4/5 VOC samples (p=0.0006). The remaining pairwise comparisons stratified by VOC obtained no significant differences (all remaining p>0.13; *Figure 5B*).

### 3.4 Determinants of RDT performance

On the condition of data availability concerning vaccination status and VOC, 1,472 of the 2,016 (73.0%) SARS-CoV-2 positive RDT/RT-qPCR tandems could be considered for the employed lasso regression model. This model revealed only the factors of viral load and typical COVID-19 symptoms being the factors directly influencing the RDT performance as both (p<0.0001) significantly increased the odds of having a positive RDT result in case of a SARS-CoV-2 infection (*Figure 5C, Supplementary Figure 6, Supplementary Table 2*).

## 4 Discussion

Overall, in a cohort of 78,798 RDT/RT-qPCR tandems, a total **sensitivity** of 34.5% was detected over the course of the COVID-19 pandemic. This places it at the lower end of the spectrum in terms of previous RDT performance analyses,[13, 47–50] consistent with interim analyses of the study.[15–18] The low sensitivity can be attributed to the study setting, which involves screening both symptomatic and asymptomatic individuals in the clinical care setting.[19] With 78,798 included RDT/RT-qPCR tandems, the presented study mapped and analysed the influence of COVID-19 vaccination and SARS-CoV-2 VOCs from the introduction and establishment of RDTs to the transition to endemicity by summer 2023 in such a large cohort in screening use including asymptomatic and symptomatic individuals. Studies of comparable size and questions only cover parts of the COVID-19 pandemic excluding consideration of Omicron BA.4/5 VOC.[19, 29]

The fluctuating number of tests throughout the study can be explained by the **RDT deployment strategy** at the study centre, which varied depending on the pandemic situation. The described RDT deployment strategy at the study centre, a tertiary care hospital, reflects the outcome of interdisciplinary development and close discussion of various interests in infection control and prevention, cost-effectiveness, and clinical feasibility.[33, 51]

Furthermore, the dominant use of MEDsan® from 2022 onwards is justified by infrastructural reasons related to cost-effectiveness and market availability. The large-scaled study cohort fairly represents the German **population** as a reference. An excess of women in their thirties can be attributed to hospitalisations related to childbirth, while the surplus of children in the first years of life can be explained a hospitalisation incidence in this age group.[46]

The conducted lasso regression analysis revealed **two significant factors directly influencing RDT performance**: exposed viral load and typical COVID-19 symptoms exhibited by the tested individual. The further univariate heterogeneities in test performance can be explained by the uneven distribution of both significant influencing factors: The lowering of RDT test performance in the chronological course of the several VOCs is caused by the obtained significant reduction of typical COVID-19 symptoms in the course of the study and therewith across the entirety of the pandemic including the chronological changes of leading VOC. The presumed decrease in sensitivity resulting from COVID-19 vaccination is also mediated by symptoms, as it was demonstrated that typical COVID-19 symptoms were significantly less commonly exhibited among vaccinated individuals.

Our findings corroborate previous reports indicating a gradual attenuation of symptoms with the evolution of VOCs in the course of the pandemic, culminating in the less symptomatic nature of the Omicron VOC, which signals the transition towards endemicity.[52–54] However, it remains undifferentiated whether the change in VOC during the course of the pandemic alone caused the change in symptoms, or whether this is also mediated by the COVID-19 specific immunity developed in the population over the course of the pandemic through previous SARS-CoV-2 infections. Further, the proportion of RDT/RT-qPCR tandems conducted on individuals vaccinated against COVID-19 significantly increased over the course of the study. Conversely, the proportion of RDT/RT-qPCR tandems involving individuals experiencing typical COVID-19 symptoms decreased derived by changes of VOC and / or immunisation status. It remains unclear whether the mitigation of symptoms during the course of the pandemic is primarily mediated by the VOC or by the immunisation status, and what proportion each of these factors contributes to the alleviation of symptoms.

It is also important to note that the increasing proportion of asymptomatic yet contagious individuals during the course of testing underscores the significance of SARS-CoV-2 screening for transmission prevention, especially when in contact with high-risk groups such as immunocompromised individuals.[55]

It is important to consider various **limitations** of the study when interpreting the data and drawing conclusions. Due to the RDT point-of-care use in the immediate clinical everyday reality in patient care and employee testing, the absolute numbers, and proportions of the RDT products used varied between different clinical departments and over the course of the study period. Similarly, reflecting the clinical care reality, the individual participating clinics of the study centre differ in the demographic patient structure and morbidity. The assignment of individual RT-qPCR methods to each sample was random based on the capacities of virus diagnostics and the clinical urgency determined by the varying processing times of different RT-qPCR methods. The study participants included were only tested with one of the three selected RDTs. Laboratory analyses assessing the test performance of RDTs in an artificial setting may provide a more comprehensive to answer the issue of comparative performance analysis of different manufacturers, but they may not be as easily translated to the population’s healthcare reality, especially with small sample sizes in the lab. The chosen RDTs in the study performed moderately in such laboratory-based performance evaluations.[13] It should also be noted that the RDTs used do not belong to the second generation of VOC-adapted RDTs, although based on the study results presented, the VOC does not directly influence the performance and therefore the first generation of RDTs can still be classified as equivalent.[56, 57] The swabs for RDT and RT-qPCR were performed by a variety of trained staff from the university hospital with user support available at all times, minimising the influence of potential heterogeneities in sample collection, test execution, and interpretation to the best extent possible. Additionally, the role of preanalytical quality and precise sample collection must be considered. Compared to RT-qPCR, RDTs are more susceptible to potentially less accurate swab collection, possibly leading to a higher rate of false-negative RDT results, especially with lower viral loads. Although the RDTs from NADAL® and MEDsan® are recommended for oro- and nasopharyngeal sampling according to the manufacturer’s instructions, the PANBIO™ RDT was also used with oropharyngeal samples, contrary to the manufacturer’s recommendations, which suggest a nasopharyngeal swab. This could limit the comparability with evidence that investigated the PANBIO™ RDT based on nasopharyngeal samples.[13] The laboratory determination of the VOC using RT-qPCR was only carried out between January 2021 and January 2022. Therefore, a relevant proportion of RDT/RT-qPCR test pairs could only be allocated to especially wild-type SARS-CoV-2 and the Omicron VOC epidemiologically. Omicron VOC sublines could only be differentiated epidemiologically with a transitional period between and before sublines BA.1-2 and BA.4-5. Allocation to other Omicron VOC sublines was epidemiologically not possible, as no other subline group in Germany exceeded the defined 90% threshold epidemiologically during the study period, and especially towards the end of the study, multiple Omicron VOC sublines were present simultaneously due to the transition to endemicity.[25] Precise sensitivity data for the Iota VOI could not be determined as only two samples detected by molecular analysis were available. In comparison to numerous published studies in the field of RDT performance analysis, the study represents a low prevalence of SARS-CoV-2 throughout the study period, with only 2.6% of included RDT/RT-qPCR test pairs showing a positive SARS-CoV-2 result.[13] However, this reflects the chosen real study setting with RDT use for SARS-CoV-2 screening, including asymptomatic test subjects. It should also be considered that vaccination data were only recorded for patients and accompanying individuals, and the recording of a COVID-19 vaccination started only from 23 May 2021. Thus, in the previous study period either no vaccination data were available, or the status “unvaccinated” could only be recorded based on age-stratified EMA approval data.[34–37, 58]

Despite all these limitations, the study’s significant value becomes apparent as it systematically examined the RDT test performance and its influencing factors in the clinical care reality over a long period of the COVID-19 pandemic, including the transition to endemicity. This includes factors that changed significantly during the pandemic - the respective dominant variants of concern (VOC) and the COVID-19 vaccination status.

In summary, this study represents a comprehensive investigation into the dynamic interaction between emerging factors such as VOC and COVID-19 vaccination throughout the entirety of the COVID-19 pandemic, utilising a substantial clinical cohort. The initially visible influence of VOC and COVID-19 vaccination could be completely mitigated identifying solely the viral load and the presence of typical COVID-19 symptoms as directly influencing factors. Both, the VOC evolution and the COVID-19 vaccinations reduced the occurrence of these symptoms in case of a SARS-CoV-2 infection and the total RDT sensitivity, but did not effect the acceptable sensitivity of RDT in typically symptomatic SARS-CoV-2 infected individuals as well as the low sensitivity in asymptomatically

infected individuals. RDTs remain a relatively reliable SARS-CoV-2 diagnostic tool in individuals showing typical respiratory symptoms even if the RT-qPCR remains the gold standard for SARS-CoV-2 diagnostics.[55, 59] Moreover, the potential of RDTs extends beyond SARS-CoV-2, as demonstrated by their adaptability to include other pathogens such as Influenza and RSV. This expansion underscores the ongoing relevance of RDTs in infection control and prevention efforts.[60, 61]

## Conflicts of Interest

Manuel Krone receives honoraria from Abbott outside of the submitted work. None of the other authors have any conflicts of interest to declare.

## Funding

This study was partially funded by the German Federal Ministry of Education and Research (BMBF) as part of the Network University Medicine (NUM): „NaFoUniMedCovid19“ Grant No: 01KX2021, Project: B-FAST as well as by COVID-19 research funds from the Free State of Bavaria provided to the Julius-Maximilians-Universität Würzburg, Germany. This work was supported by Bay-VOC (Bavarian State Ministry of Health and Care). Nils Petri was supported by the UNION CVD scholarship funded by the German Research Foundation (DFG).

## Role of the funding source

This study was initiated by the researchers themselves. The funding institutions had no influence on the study design, data collection, analysis and interpretation, or the writing of the manuscript. All authors had unrestricted access to all data. The first and corresponding authors were responsible for the final decision to submit the study for publication.

## Data Access

Requests for access to an anonymised version of the complete dataset underlying this analysis can be made to the corresponding author with a specific question, for a period of up to 5 years from the immediate time of publication.

## Data Availability

Requests for access to an anonymized version of the complete dataset underlying this analysis can be made to the corresponding author with a specific question, for a period of up to 5 years from the immediate time of publication.

## Acknowledgments

We would like to thank all involved employees of the University Hospital Würzburg for the realisation and careful documentation of the RDTs as well as the establishment of the RDT infrastructure.

Professor Ulrich Vogel was significantly involved in the conception and design of the study, as well as in the procurement of funds, user support, and assistance. Unfortunately, he passed away during the study period in 2022, and was unable to review and approve this manuscript. We miss him as an enthusiastic colleague and friend who was deeply committed to his work, family, and friends.

## Contributions

All authors had unlimited access to all data. Isabell Wagenhäuser and Manuel Krone take responsibility for the integrity of the data and the accuracy of the data analysis.

**Conceptualisation**: Isabell Wagenhäuser; Kerstin Knies; Ralf-Ingo Ernestus; Johannes Forster; Dirk Weismann; Benedikt Weißbrich; Johannes Liese; Christoph Härtel; Oliver Kurzai; Lars Dölken; Alexander Gabel; Manuel Krone

**Methodology**: Isabell Wagenhäuser; Kerstin Knies; Alexander Gabel; Manuel Krone

**Software**: Isabell Wagenhäuser; Alexander Gabel; Manuel Krone

**Validation**: Isabell Wagenhäuser; Kerstin Knies; Tamara Pscheidl; Michael Eisenmann; Sven Flemming; Nils Petri; Miriam McDonogh; Agmal Scherzad; Daniel Zeller; Anja Gesierich; Anna Katharina Seitz, Regina Taurines; Ralf-Ingo Ernestus; Johannes Forster; Dirk Weismann; Benedikt Weißbrich; Johannes Liese; Christoph Härtel; Oliver Kurzai; Lars Dölken; Alexander Gabel; Manuel Krone

**Formal analysis**: Isabell Wagenhäuser; Alexander Gabel; Manuel Krone

**Investigation**: Isabell Wagenhäuser; Kerstin Knies; Tamara Pscheidl; Michael Eisenmann; Sven Flemming; Nils Petri; Miriam McDonogh; Agmal Scherzad; Daniel Zeller; Anja Gesierich; Anna Katharina Seitz; Regina Taurines

**Resources**: Isabell Wagenhäuser; Kerstin Knies; Tamara Pscheidl; Michael Eisenmann; Sven Flemming; Nils Petri; Miriam McDonogh; Agmal Scherzad; Daniel Zeller; Anja Gesierich; Anna Katharina Seitz; Regina Taurines; Ralf-Ingo Ernestus; Dirk Weismann; Benedikt Weißbrich; Johannes Liese; Christoph Härtel; Lars Dölken

**Data curation**: Isabell Wagenhäuser; Kerstin Knies; Tamara Pscheidl; Manuel Krone

**Writing - Original Draft**: Isabell Wagenhäuser; Manuel Krone

**Writing - Review & Editing**: Kerstin Knies; Tamara Pscheidl; Michael Eisenmann; Sven Flemming; Nils Petri; Miriam McDonogh; Agmal Scherzad; Daniel Zeller; Anja Gesierich; Anna Katharina Seitz; Regina Taurines; Ralf-Ingo Ernestus; Johannes Forster; Dirk Weismann; Benedikt Weißbrich; Johannes Liese; Christoph Härtel; Oliver Kurzai; Lars Dölken; Alexander Gabel

**Visualisation**: Isabell Wagenhäuser; Alexander Gabel; Manuel Krone

**Supervision**: Alexander Gabel; Manuel Krone

**Project administration**: Isabell Wagenhäuser; Kerstin Knies; Alexander Gabel; Manuel Krone

**Funding acquisition**: Oliver Kurzai; Lars Dölken

The manuscript was reviewed and approved by all authors.

## Statement on the use of artificial intelligence (AI) in the writing process

Artificial intelligence (AI) was used for language improvement purposes. The tools ChatGPT (OpenAI, San Francisco CA, USA) and DeepL (DeepL SE, Cologne, Germany) were used. The actual writing of the manuscript was carried out solely by the authors mentioned by name.

## Information on previous data presentation

Four studies have already been conducted as interim analyses in advance including subsets and sub-populations of the overall data set used for this study. These include two analyses of age independent RDT performance within a cohort of 5,068 individuals. Data collection for these studies spanned from the study commencement on 12 November 2020, dduntil 28 February 2021, and continued until 30 January 2022. In total, 35,479 RDTs were administered during this period, without accounting for BA.4/5 VOC.[15, 16] The analysis specific to the paediatric population, covering the data collection period from 12 November 2020, to 30 September 30 2022, was conducted separately and subsequently published.[17] A further brief evaluation of 54,740 parallel RDT/RT-qPCR oropharyngeal swabs of 38,373 adults ≥ 18 years of age from the same period, which chronologically considered all SARS-CoV-2 VOC up to and including Omicron BA.4/5 without consideration of COVID-19 vaccination status, has also been published.[18]

## Data sharing statement

Additional data that underlie the results reported in this article, after de-identification (text, tables, figures, and appendices) as well as the study protocol, statistical analysis plan, and analytic code is made available to researchers who provide a methodologically sound proposal to achieve aims in the approved proposal on request to the corresponding author.

