## Supplementary Material for "SARS-CoV-2 Antigen Rapid Detection Tests: test performance during the COVID-19 pandemic and the impact of COVID-19 vaccination"

#### Supplementary Methods

##### Determination and allocation of SARS-CoV-2 virus variant of concern

Between 3 February 2021, and 19 January 2022, all RT-qPCR-positive samples with sufficient viral load underwent PCR with spike protein variant-specific primers to differentiate between current VOC using the VirSNiP SARS-CoV-2 Spike N501Y, del 69/70, E484K, N501Y, L452R, T478K, and 371L 373P 452R Kits (TIB molbiol, Berlin, Germany) carried out on a cobas z 480 analyzer (Roche Diagnostics, Rotkreuz, Switzerland). This method was chosen because VOC differentiation through multiple RT-qPCR tests showed reliable and high agreement with VOC determination using SARS-CoV-2 genome sequencing. In the case of combinations of results from this method that did not correspond to a common VOC, the described analytical procedure was carried out using SARS-CoV-2 Spike protein sequencing to molecularly determine SARS-CoV-2.[1] With very high COVID-19 incidence in the pandemic phase with almost complete dominance of the Omicron VOC at very high RT-qPCR sample volume, the laboratory analytical VOC determination was finally terminated on 19 January 19 2022.[2]

For all RT-qPCR samples collected outside the period of molecular VOC determination, the VOC was epidemiologically assigned to the RT-qPCR sample. For RT-qPCR samples collected within the period of laboratory VOC determination but with low viral load making molecular VOC determination not possible or in VOC determination, no VOC could be clearly diagnosed; if available, the VOC was derived from the known SARS-CoV-2 infection source; otherwise, the VOC was similarly epidemiologically assigned. This was done according to the following principle: All RT-qPCR samples included before 3 February 2021, were assumed to be SARS-CoV-2 wild-type since, chronologically according to epidemiology, the first VOC detections in Germany occurred after this date. Epidemiologically, the RT-qPCR sample was always assigned to the VOC responsible for more than 90% of SARS-CoV-2 cases in Germany in the calendar week of sample collection. If no VOC in a calendar week caused more than 90% of SARS-CoV-2 cases, no VOC assignment to a RT-qPCR positive sample was possible; those intervals were referred to as the "transition period".[3]

With the spread of the Omicron VOC, there was also the epidemiological peculiarity that various Omicron VOC sublines occurred during the entire Omicron VOC-caused phase of the COVID-19 pandemic, with the associated transition to the endemic phase. From the third calendar week of 2022 until the end of the study, epidemiologically, in total, at least 90% of SARS-CoV-2 cases were caused by different Omicron VOC sublines. Since only for the two Omicron VOC subline cohorts Omicron BA.1-2 VOC and Omicron BA.4-5 VOC, a clear VOC assignment could be defined by reaching the defined epidemiological threshold of 90% during the respective periods, outside of this period from the third calendar week of 2022, an "Omicron VOC transition period" was defined where parallel different sublines were detectable, making up more than 90% of SARS-CoV-2 cases, but no single VOC or VOI reached this threshold. If in the molecular analysis, the VOC was only differentiated as Omicron VOC, in the case of an analysis stratified according to the Omicron VOC sublines BA.1-2 and BA.4-5, the epidemiological allocation of the molecular result was preferred as an exception (*Supplementary Table 1*).[3]

### Supplementary Tables and Figures

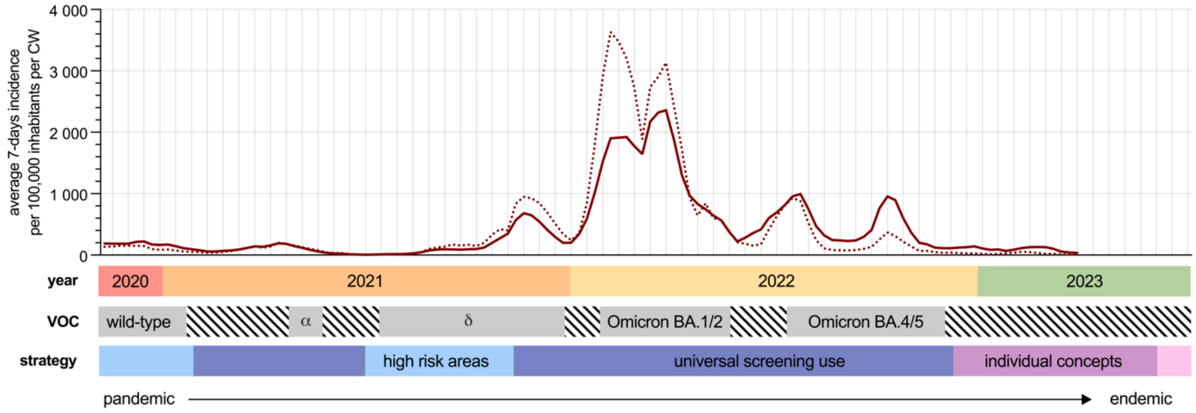


**Supplementary Figure 1**: SARS-CoV-2 incidence in Bavaria during the COVID-19 pandemic, the SARS-CoV-2 VOCs and the RDT deployment strategy of the study

The solid curve shows the incidence in the general population, the dashed curve the incidence among children and adolescents aged 0-15 years. The data was not available from the provider until the end of the study.

VOC: virus variant of concern

Data source: Robert Koch-Institut, Bayerisches Landesamt für Gesundheit und Lebensmittelsicherheit[2, 3]

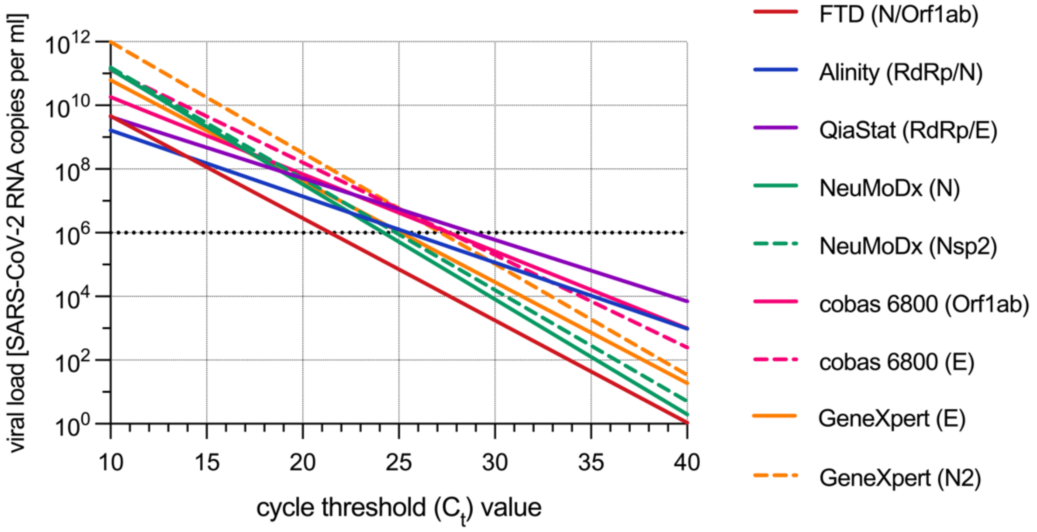


**Supplementary Figure 2**: Viral load as a function of the $C_t$ value stratified according to the RT-qPCR method

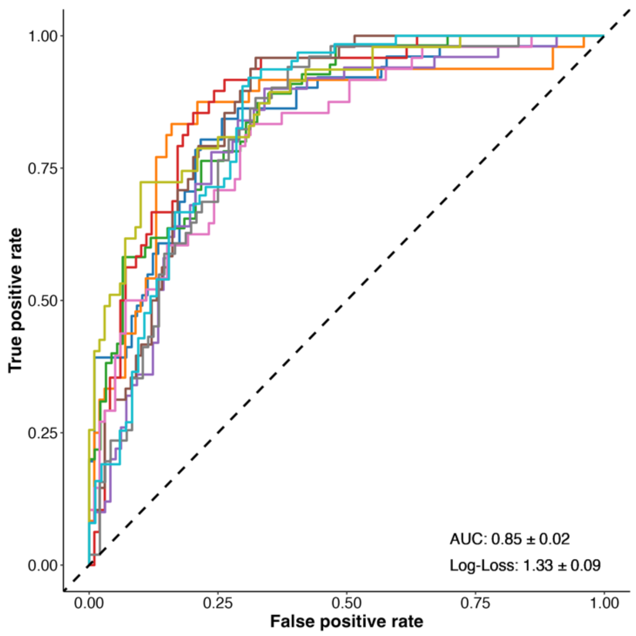


**Supplementary Figure 3**: Lasso regression model: AUC (area under the curve) of the ROC (receiver operating characteristic) curve

| calendar week (CW) | epidemiological VOC-allocation |
|---|---|
| CW 46/2020 - CW 3/2021 | SARS-CoV-2 wild-type |
| CW 4/2021 - CW 15/2021 | *VOC transition* |
| CW 16/2021 - CW 17/2021 | Alpha VOC |
| CW 18/2021 - CW 19/2021 | *VOC transition* |
| CW 20/2021 - CW 21/2021 | Alpha VOC |
| CW 22/2021 - CW 27/2021 | *VOC transition* |
| CW 28/2021 - CW 50/2021 | Delta VOC |
| CW 51/2021 - CW 2/2022 | *VOC transition* |
| CW 3/2022 - CW 20/2022 | Omicron BA.1-2 VOC |
| CW 21/2022 - CW 26/2022 | *Omicron VOC transition* |
| CW 27/2022 - CW 47/2022 | Omicron BA.4-5 VOC |
| CW 48/2022 - CW 26/2023 | *Omicron VOC transition* |

**Supplementary Table 1**: Epidemiological VOC assignment by calendar week (CW) in the study period

VOC: virus variant of concern

Data source: Robert Koch-Institut[3]

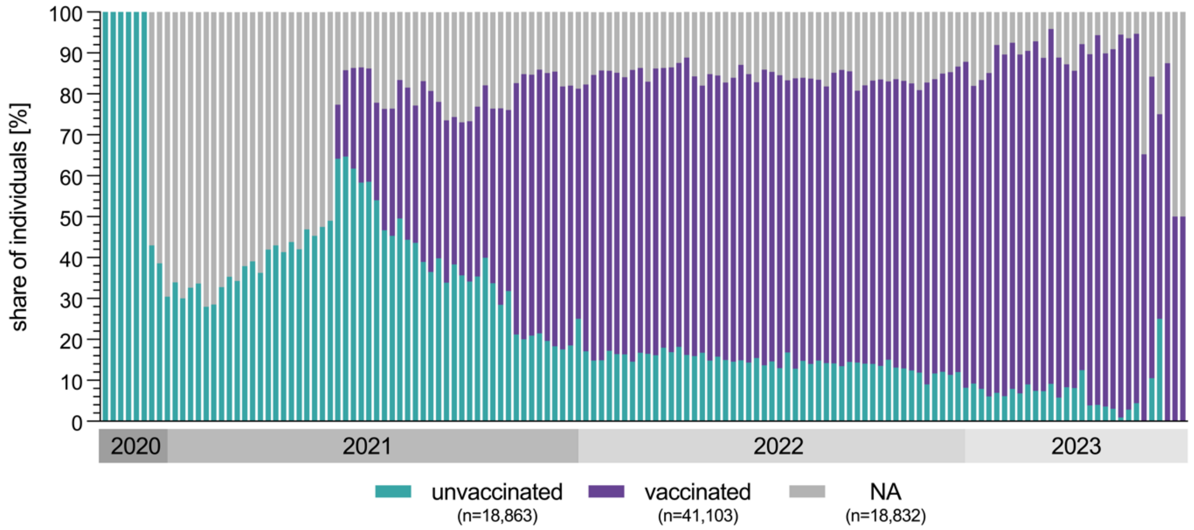


**Supplementary Figure 4**: Fraction of COVID-19 vaccination status of RDT/RT-qPCR test tandems per CW including individuals with no available information on COVID-19 vaccination status

NA: no information available

Data source: European Medicines Agency (EMA)[4-7]

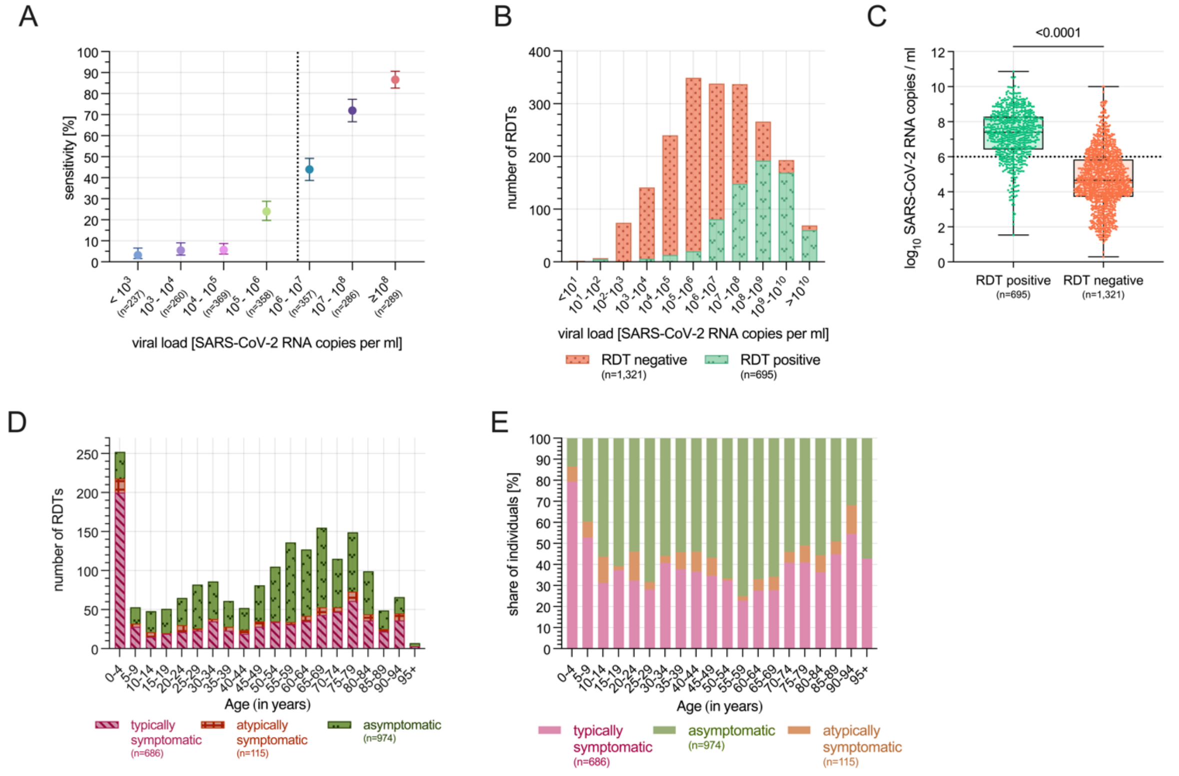


**Supplementary Figure54**: Distribution of potential RDT performance influencing factors

5A) RDT sensitivity depending on viral load in categories

5B) RDT test result depending on viral load in absolute numbers

5C) Viral load stratified by RDT test result

5D) Age distribution of RDTs stratified by COVID-19 symptomatology

5E) Proportionate distribution of symptomatology stratified by age category

RDT: Antigen Rapid Detection Test

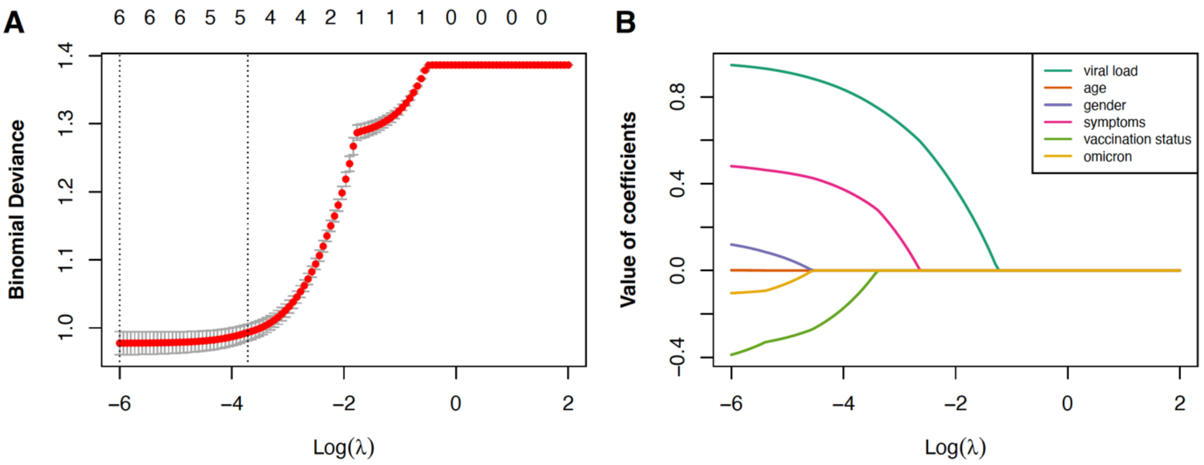


**Supplementary Figure 6**: Lasso regression for detection of influencing factors associated to RDT sensitivity

6A) Tenfold cross-validation procedure to determine optimal lambda parameter based on minimal mean-squared error

6B) Illustrating the shrinkage of coefficients (factors) towards zero with increasing lambda values

| | number RT-qPCR positive | positive RDT result | sensitivity | OR univariate | OR multivariate |
|---|---|---|---|---|---|
| **ordinate variables** | | | | | |
| *age* | - | - | - | - | 1.00 (0.99-1.01) |
| *viral load* | - | - | - | - | 2.61 (2.34-2.90) |
| **gender** | | | | | |
| *female* | 976 (48.4%) | 346 | 35.5 (32.5-38.5) | 0.55 (0.23-1.32) | 1.16 (0.87-1.54) |
| *male* | 1,040 (51.6%) | 349 | 33.6 (30.8-36.5) | 0.51 (0.21-1.22) | - |
| **COVID-19 symptoms** | | | | | |
| *typically symptomatic* | 686 | 359 | 52.3 (48.6-56.1) | 1.10 (0.80-1.50) | 2.45 (1.79-3.36) |
| *atypically symptomatic* | 115 | 52 | 45.2 (36.4-54.3) | 0.83 (0.53-1.32) | 1.4 (0.79-2.54) |
| *asymptomatic* | 974 | 221 | 22.7 (21.2-25.4) | 0.29 (0.21-0.40) | - |
| *no symptom information* | 275 | 80 | 29.1 (24.0-34.7) | - | - |
| **COVID-19 vaccination status** | | | | | |
| *vaccinated* | 1,091 | 333 | 30.5 (27.9-33.3) | 0.44 (0.32-0.60) | 0.91 (0.59-1.40) |
| *unvaccinated* | 525 | 217 | 41.33 (37.2-45.6) | 0.70 (0.51-0.97) | - |
| *no vaccination information* | 400 | 145 | 36.3 (31.7-41.1) | - | - |

**Supplementary Table 2**: RDT sensitivity and its potential influencing factors
OR: Odds Ratio
