## Supplementary figures and images for "SARS-CoV-2 Antigen Rapid Detection Tests: test performance during the COVID-19 pandemic and the impact of COVID-19 vaccination"

### Supplementary Figure 1

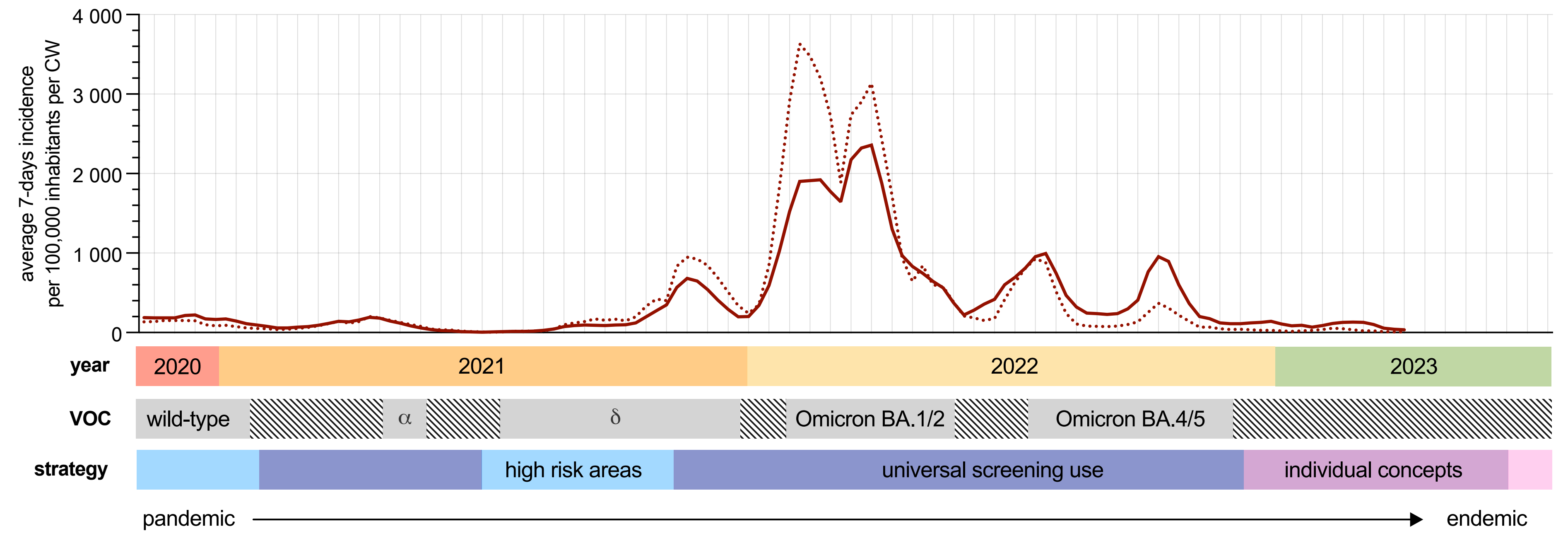

### Supplementary Figure 2

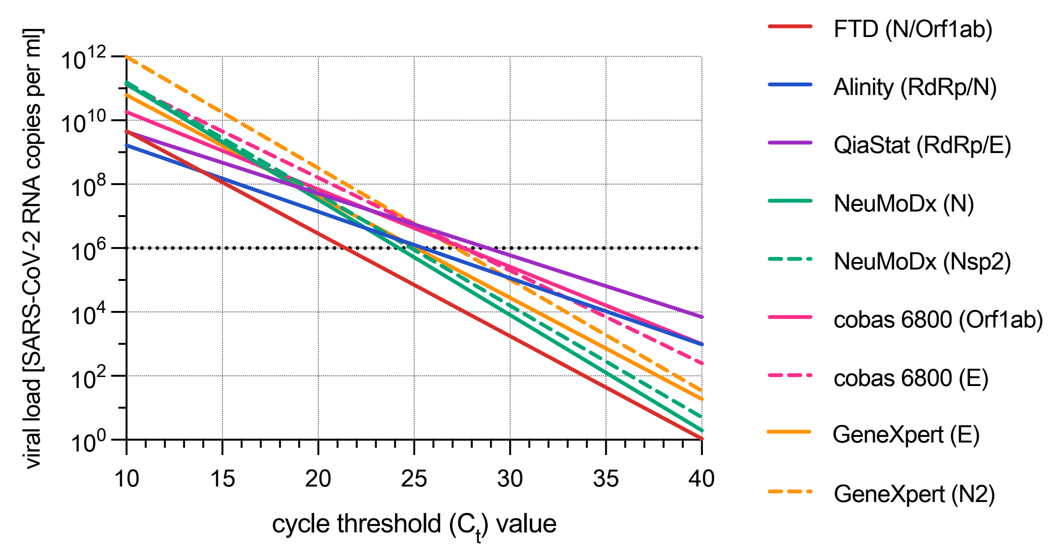

### Supplementary Figure 3

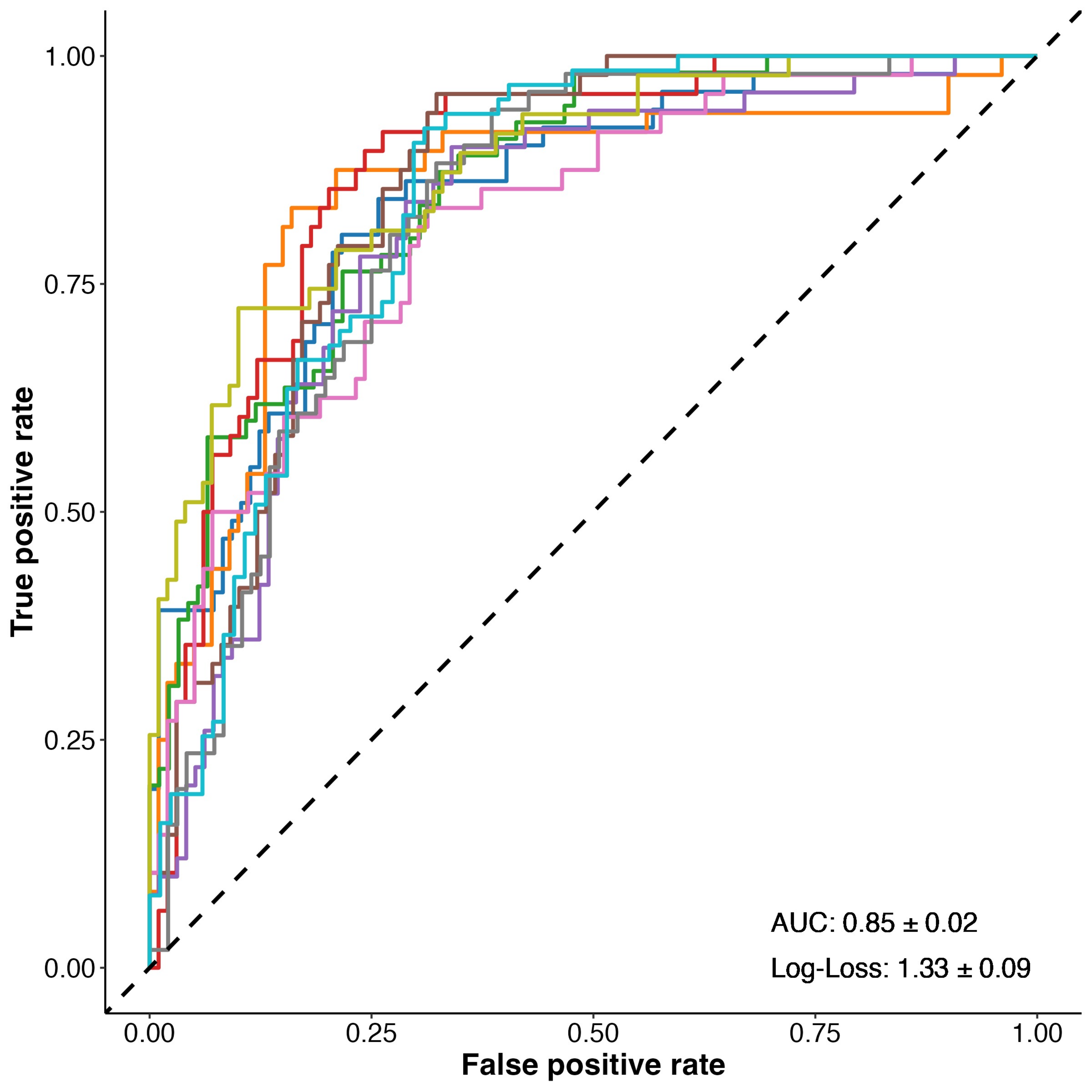

### Supplementary Figure 4

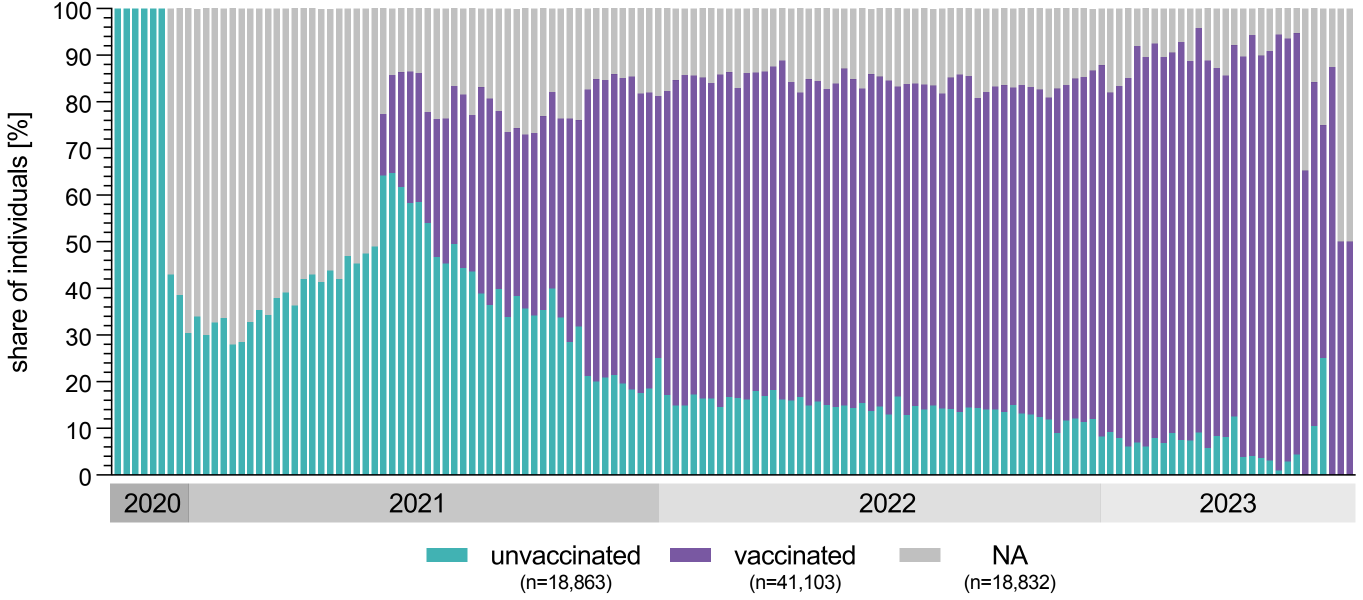

### Supplementary Figure 5

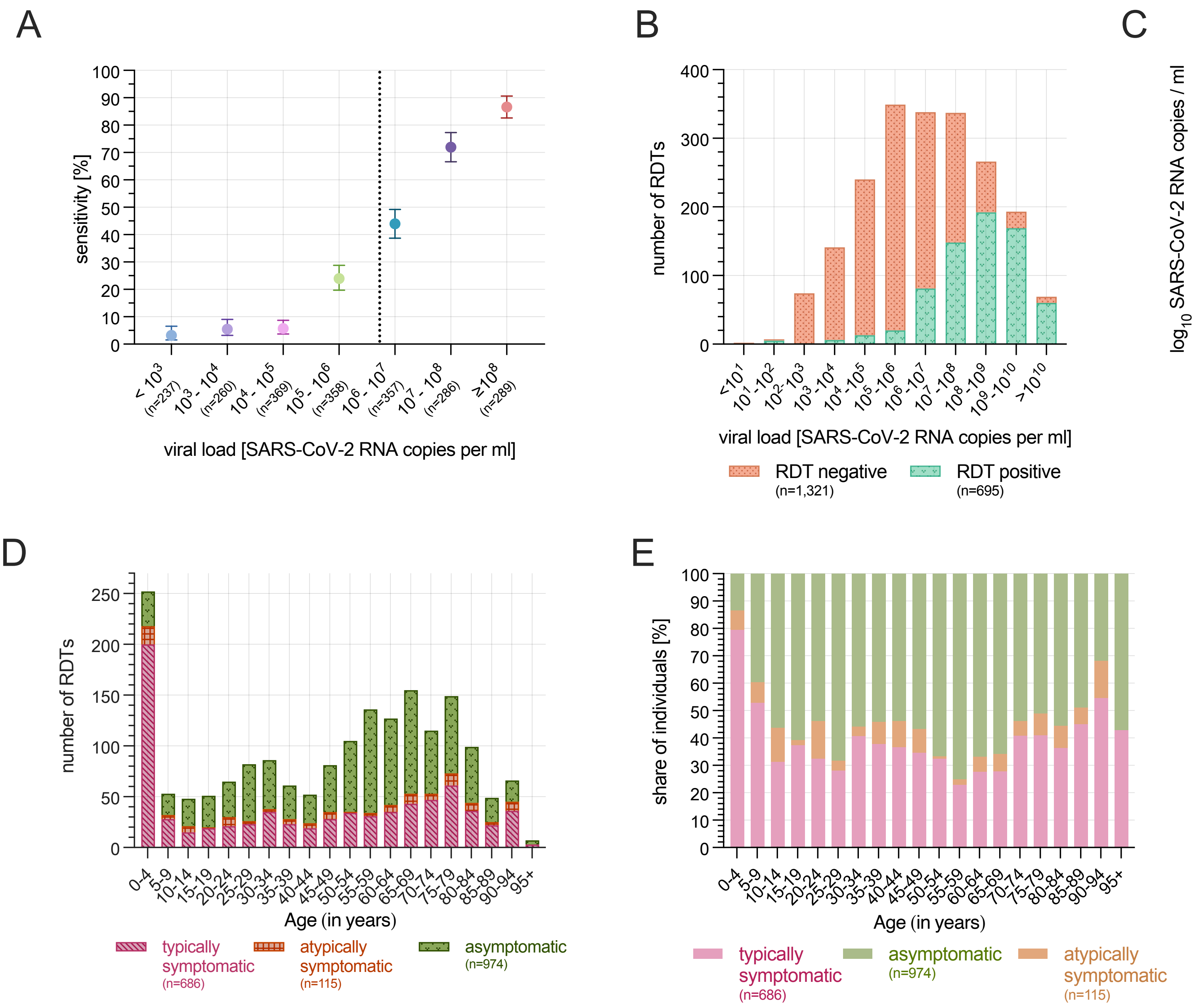

### Supplementary Figure 6

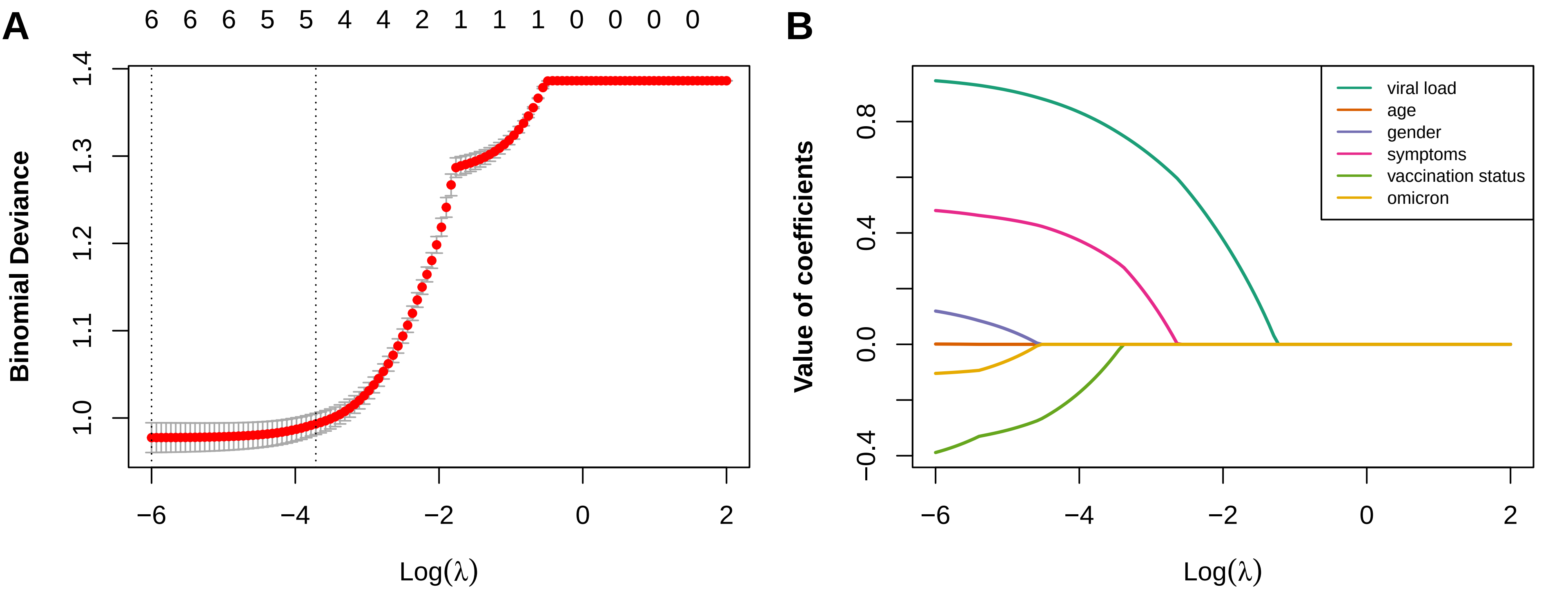
